# Digital Health Interventions (DHIs) for Health Systems Strengthening in Sub-Saharan Africa: Insights from Ethiopia, Ghana, and Zimbabwe

**DOI:** 10.1101/2025.04.22.25326213

**Authors:** T Simbini, E Adimado, S Adjorlolo, L Guerrero-Torres, P N Srinivas, S Zizhou, TA Zerfu

**Author notes:** Authors contributed equally in this work.

## Abstract

Digital health interventions (DHIs) refer to discrete technological functionalities designed to achieve specific objectives in addressing health system challenges through digital health applications. These interventions are tools for strengthening health systems, particularly in low- and middle-income countries. This study consolidates findings from Ethiopia, Ghana, and Zimbabwe, examining three distinct digital health applications with varying intervention capabilities that contribute to health system strengthening within their respective primary healthcare contexts. The interventions analyzed include Ethiopia’s District Health Information System 2 (DHIS2), Ghana’s dual system namely the District Health Information Management System (DHIMS) and the Local Health Information Management System (LHIMS and Zimbabwe’s *Impilo* electronic health record (E-HR) system.

In Ethiopia, DHIS2 supports DHIs focused on data aggregation, reporting, and performance monitoring at the public health level. At the health system level, DHIS2 has enhanced accountability and data quality, leading to improved decision-making and resource distribution. In Ghana, DHIMS functions as a public health-level DHI, facilitating data-driven performance monitoring, while LHIMS operates at the patient level, supporting patient tracking and management, improving patient workflows and resource tracking. However, a lack of interoperability between these two systems has led to data duplication challenges. Zimbabwe’s *Impilo* E-HR, a patient-level DHI, has streamlined clinical workflows, improved information sharing, and enhanced decision-making at the point of care.

Despite these successes, challenges persist, including infrastructure limitations, high staff turnover, and insufficient technical capacity among users. Interoperability issues, particularly in Ghana and Ethiopia, hinder seamless data exchange, while sustainability concerns such as funding gaps and inadequate government support limit the full potential of these systems. The studies highlight the need for targeted training, improved infrastructure, and enhanced integration of health information systems to maximize the benefits of DHIs.

**Authors’ Summary:** Health systems are built on six fundamental building blocks, yet they face numerous challenges, particularly in the African context. Digital health interventions (DHIs) have been shown to strengthen one or more of these building blocks by improving healthcare efficiency, data management, and service delivery. These interventions are especially valuable in low- and middle-income countries, where they help address systemic issues such as inefficient data management, limited access to care, and health workforce efficiencies.

Our paper presents findings from three independent studies examining digital health applications implemented in Ethiopia, Ghana, and Zimbabwe, each playing a unique role in strengthening their respective health systems. We explore how DHIs have contributed to key health system functions, highlighting successes in data utilization, patient management, and decision-making. At the same time, we report on the challenges faced by implementers, including infrastructure limitations, interoperability issues, and sustainability concerns.

Based on our analysis and experience of the environment, we offer insights and practical recommendations for overcoming these challenges, ensuring that DHIs can maximize their impact on healthcare delivery. By addressing these barriers, DHIs have the potential to enhance health system resilience and efficiency, ultimately improving health outcomes in resource-limited settings.

## Introduction

Digital health interventions (DHIs) refer to discrete technological functionalities designed to address specific health objectives through digital health applications (1). As implemented across various digital health applications, DHIs are increasingly recognized as pivotal tools for strengthening health systems in low- and middle-income countries. In Sub-Saharan Africa (SSA), where healthcare systems often grapple with limited resources, fragmented services, and logistical challenges, digital technologies offer innovative solutions to enhance service delivery and health outcomes (2). The adoption of digital health tools in these countries has led to significant improvements in various aspects of health systems. For instance, electronic health records and mobile health applications have facilitated better data management, improved patient tracking, and enhanced communication between healthcare providers. These technologies have enabled more efficient health service delivery, contributing to better patient outcomes and more robust health systems. DHIs, implemented through digital health applications, enhance health system functionality at various levels. They support improved public health reporting through electronic aggregate reporting tools such as District Health Information System 2 (DHIS2) and facilitate better patient tracking using electronic medical record systems. By integrating digital health interventions (DHIs), countries have been able to address some of the critical gaps in their healthcare infrastructure, making strides towards more effective and equitable health systems (3). Despite the promising advancements, the implementation of DHIs in Sub-Saharan Africa is fraught with challenges. Inadequate infrastructure, limited technological literacy, and data privacy concerns often hinder the effectiveness of these initiatives. Additionally, the sustainability of digital health projects is a recurring challenge, with many interventions facing difficulties in maintaining financial and operational support over the long term. (4,5). These barriers can impede the full realization of the potential benefits of digital health technologies, necessitating a closer examination of their practical limitations and strategies for overcoming them.

In this paper, we consolidate three country studies that investigated the role of DHIs in strengthening health systems in Ethiopia, Ghana, and Zimbabwe, with a focus on primary health care. The study from Ethiopia evaluated the District Health Information System 2 (DHIS2) with respect to accountability, accessibility, equity, and data utilization in primary health facilities. The Ghana study assessed the effectiveness of two systems the District Health Information Management System (DHIMS), a repository of national aggregate data on critical health indicators, and the Lightwave Health Information Management System (LHMIS), a local electronic health record system, on how they improved access to and use of health data by primary health managers. The Zimbabwe study examined the contribution of the *Impilo* electronic health record system on health systems strengthening, emphasizing improvements in care quality and health equity at the primary care level. Therefore, this paper aims to explore common opportunities and challenges associated with DHIs in strengthening health systems in Ghana, Ethiopia and Zimbabwe. It provides insights into how various DHIs influence health service delivery, data management, and overall system effectiveness, offering guidance for future implementations and improvements in similar settings.

## DHIs Implementation Context

### Ethiopia

Ethiopia has made remarkable strides in incorporating digital technologies into its healthcare system, with a particular focus on reaching rural and underserved areas. The country’s diverse geographic and socio-economic environments have provided essential insights into how digital health solutions can be effectively adapted to different contexts, ranging from urban centers to remote villages. Since 2017, Ethiopia has implemented DHIS 2 as its standard platform for data collection and reporting, which is now successfully operating in over 4,000 healthcare facilities across the nation, improving healthcare delivery and monitoring.

### Ghana

By 2017, Ghana had implemented two digital health information systems namely the District Health Information System (DHIS2), locally named the District Health Information Management System (DHIMS), which serves as a repository for aggregate data for the largest implementing agency within the health sector, the Ghana Health Service (GHS) and an electronic health management system known as Lightwave Health Information Management System (LHIMS) hosted by the Ministry of Health. The DHMIS is deployed in all the primary and secondary healthcare facilities within the GHS while the LHIMS is deployed in the country’s district, regional hospitals, and teaching hospitals. These two systems are currently not interoperable. The target users of both systems are all healthcare professionals and policy makers within the health sector. Currently, most public health facilities within the 16 regional capitals are using the LHIMS, while efforts are ongoing to extend to other points of care like the health centers.

### Zimbabwe

Zimbabwe adopted digital health interventions (DHIs) for patient tracking in 2013, initially focusing on monitoring patients on antiretroviral therapy (ART) (6). In 2016, the country piloted *Impilo*, a locally developed web-based electronic health record (E-HR) system, to expand patient tracking beyond HIV/AIDS treatment. By 2023, *Impilo* had become the primary E-HR system in public health facilities, implemented in 1,055 of the 1,800 primary and secondary care facilities nationwide. The system includes 18 modules covering various clinical care departments, such as Patient Registration, General Outpatients Consultation, HIV/AIDS/TB/STIs, Pharmacy and Commodity Tracking (7), effectively supporting all health system building blocks at these levels of care. The system is built on the Open Health Information Exchange (Open HIE) framework and integrates Health Level 7 Fast Interoperability Resources (HL7 FHIR), enabling seamless interoperability with other systems, including laboratory information management systems and automated reporting in DHIS2 (8).

## Research Methods

The overall study consisted of three mixed methods research studies at each site followed by an analysis of findings across the three sites.

**Ethiopia** conducted a mixed-methods study across five regions to evaluate the impact of DHIS2 on maternal and child health outcomes. The overarching goal of the study was to evaluate how DHIS2 has influenced data availability, accessibility, and decision-making in Ethiopia’s health system. The findings aim to identify the strengths and challenges of DHIS2, ultimately guiding improvements in health data management and ensuring better health outcomes for mothers and children across the country. The study used Key Informant Interviews (KIIs) and Focus Group Discussions (FGDs) with participants from Primary Health Care Units (PHCUs) across five regions. These provided valuable insights into the perspectives of health workers and community members regarding the use and effectiveness of DHIS2 in improving health services. In addition to the qualitative data, the study also reviewed performance reports from 2013 to 2022, alongside electronic health data, to assess the impact on the system over time.

The quantitative data were analysed using Interrupted Time Series Analysis (ITSA), which helped identify trends and patterns in maternal and child health indicators before and after the implementation of DHIS2. The qualitative data were thematically coded to extract key insights on the accessibility, quality, and equity implications of DHIS2.

**Ghana** conducted a sequential mixed-methods study to assess how two digital health interventions, DHIMS and LHIMS, enhance access to and use of healthcare data by primary healthcare (PHC) managers. The study involved 125 PHC managers from 10 district hospitals and one regional hospital in the Greater Accra Region. The first phase involved the administration of survey questionnaires using cross-sectional design to generate quantitative summaries of the application of DHIMS and LHIMS from hospital unit managers/in-charges. This was followed by a qualitative exploratory approach involving individual in-depth interviews (IDIs) to obtain detailed and in-depth insights into the DHIMS and LHIMS from PHC managers. Both quantitative and qualitative data were triangulated to obtain richer description and understanding of the application of DHIMS/LHIMS. The research focused on the contributions of DHIMS and LHIMS in data utilization, examined interoperability issues between the systems, and identified challenges in using the healthcare data provided by these platforms.

**Zimbabwe** conducted a quasi-experimental, non-randomized mixed-methods study to assess the impact of E-HR on health systems building blocks, quality of care, and implementation challenges in resource-limited settings. The study was conducted in two purposively sampled provinces. In each province, an E-HR implementing district and a non-implementing district were selected. The selected implementing districts began using E-HR in 2021. Thirty-two health facilities were sampled, twenty (20) from E-HR implementing sites and twelve (12) from non-implementing sites. District hospitals were automatically included. The study population were 21 key informants (district health executive), 37 health care providers (nurses, doctors, pharmacists and laboratory scientists) and 173 clients receiving care. These were interviewed as key informants. The study investigated E-HR contributions on each of the health system building blocks, quality of care, and the role of the DHIs in improving decision-making, equity, quality, and efficiency, while documenting lessons from large-scale E-HR deployment.

## Ethical Approvals

### Ethiopia

Ethical approval was obtained from five regional Institutional Review Boards (IRBs) under the respective health bureaus: Addis Ababa City Administration and the four regions (Oromia, Amhara, Sidama, and Somali). **Ghana.** The study received ethical approval from the Ghana Health Service Ethical Review Committee (GHS-ERC003/04/23).

### Zimbabwe

This study was approved by the Joint Research and Ethics Committee JREC at the University of Zimbabwe, the Ministry of Health through the Permanent Secretary and the Medical Research Council of Zimbabwe under study ID MRCZ/A/3127.

## Ethics Statement

All study participants in Ethiopia, Ghana, and Zimbabwe provided verbal informed consent prior to their participation. The purpose of the study, the voluntary nature of participation, confidentiality measures, and the right to withdraw at any time were explained to each participant. Verbal consent was recorded at the beginning of each interview, in accordance with ethical guidelines for studies involving minimal risk and where written consent was not feasible.

## Results

The three studies investigated the digital health information systems and their contributions to one or more building blocks of health systems. We categorize the findings from the three countries based on the applicable health systems building blocks.

### Health Information Systems

Findings from the three studies reported the increased usage of these tools at different levels of the health system. Primary and secondary care facilities have largely adopted these tools in all settings. For all three country settings, aggregate health data reporting is based on DHIS2 and all primary and secondary facilities are reporting in DHIS2.

The Ethiopian study found that the District Health Information System 2 (DHIS2) plays a crucial role in several key healthcare functions. Of the facilities that reported using DHIS2, 99% use it for reporting, 96% for program monitoring, 67% for target setting, and 87% for resource allocation. These functions have contributed to improving the management of healthcare services, ensuring that resources are allocated effectively to areas that need them most. DHIS2 has helped streamline the reporting processes, making it easier for healthcare providers and administrators to monitor progress and identify areas requiring attention. This has been particularly important in making evidence-based decisions to improve service delivery across various health programs. Patient level information systems are seeing an increased uptake in both Ghana and Zimbabwe. In Ghana, all primary and secondary facilities have adopted the LHIMS. In Zimbabwe, 56% of primary and secondary facilities have adopted E-HR. Data is cascaded from facility to national level in all cases. There is increased reliance on these digital health information management systems by service providers, and decision makers. In Ethiopia, DHIS2 has significantly improved data quality, availability, accessibility, and accountability by streamlining data collection and enabling real-time reporting. It has ensured consistent and accurate reporting across health facilities, reducing errors and delays. The system has also enhanced accessibility for policymakers, allowing for better resource allocation and decision-making. Additionally, DHIS2 has promoted accountability by tracking performance and ensuring that data informs public health strategies, ultimately strengthening the overall healthcare system. Similarly, in Ghana, both DHIMS and LHIMS have been integral to data-driven decision-making for primary healthcare (PHC) managers, particularly in forecasting, monitoring, and evaluating performance. In Zimbabwe, the electronic health records (E-HR) has improved information sharing among health practitioners, enhanced data integrity through audit trails, and strengthened decision support at the point of care, as note by the following responses:

> *“Increased credibility in data, what you enter into the E-HR is what is displayed in the E-HR unlike when using paper” [Zim Key Informant]*

Accessing patient level information was also noted to improve:

> “*Reduces time taken to find the patient’s record*” [Zim Nurse Manager]

It has also streamlined national reporting requirements reducing the number of manual aggregate reports and registers generated, hence leaving more time available to patient care:

> “*It can improve workforce effectiveness…, no more registers required to be written which consumes most of the time.”* [Zim Key Informant]

Integration of E-HR with DHIS2 has reduced the reporting requirements on nurses and increased the reliability and timeliness of reported data.

### Service Delivery

Ethiopia and Zimbabwe both reported improvements in service delivery due to their respective DHIs. In Zimbabwe, the E-HR system streamlined clinical workflows, particularly in prescriptions, reducing patient wait times and increasing the efficiency of service delivery.

> *"E-HR ’Impilo’ has streamlined our processes and improved overall service delivery."* [Key Informant]

The availability of real-time patient records has enhanced patient experiences by minimizing health care worker time spent on documentation tasks. However, not all users share the same sentiments, some users noted that using the E-HR system became more time-consuming during periods of high patient volumes. The slow response times of these applications, during high patient volume reduces the effectiveness of E-HR at point of care:

> “*The way I see it, it’s a burden. In the case of a patient who comes for HIV testing, it takes 5 to 10 minutes to read the records and yet the patients are waiting in the queue, we end up resorting to shortcuts because I still have to attend to others….* [Zim Health Worker].

In Ethiopia, DHIS2 has similarly contributed to more efficient healthcare delivery by improving data accuracy and timeliness, which are crucial for effective resource allocation and service planning. For instance, a PHCU head from one region highlighted the significant improvement in timeliness after DHIS2 implementation, saying:

> "*Before DHIS2, we struggled with tracking data timeliness. After implementation, the transformation was clear - our data management reports are now more timely than ever."* [Eth PHCU head]

### Health Workforce

All three countries have reported the increased use and integration of systems within the various service departments including clinical care workflows. The implementation of DHIS2 in Ethiopia and Ghana has significantly improved accountability within the healthcare sector. This system enables policymakers at district/regional and national levels to monitor resource utilization and assess the impact of healthcare interventions more effectively.

However, challenges related to technical capacity have emerged as a significant barrier to the full utilization of DHIS2. A shortage of adequately trained personnel in many primary healthcare facilities across all three countries was noted, hindering optimal system operation. Even among trained staff, the lack of continuous technical support exacerbates these challenges. Furthermore, Zimbabwe and Ethiopia reported high staff turnover for those trained in the use of the DHIs. These limitations in technical proficiency and support negatively affect the efficiency with which the system is utilized, leading to potential inaccuracies and incompleteness in data collection. Consequently, the system’s full potential to enhance health information management and improve decision-making remains underutilized. Ghana noted 88% of interviewed PHC managers relied on LHIMS data, while 70% of them obtained summary data from DHIMS. About 73% reported that the DHIMS/LHIMS was easy/very easy to access and majority of the participants, 86% indicated that the DHIMS was presented in an easy-to-understand format, compared with the LHIMS (63%). More importantly, about 96% (n = 88) and 81% (n = 95) noted the information in DHIMS and LHIMS to be *Very Useful* and *Useful*, respectively.

> “*It helps us to make decisions. For example, if I know the number of deliveries that I have had for a period it will help me to improve my system. When I say system, I mean both managerial as well as logistical*” (Gha-PHC manager)

Ghana’s experience highlighted the struggle of older healthcare professionals, particularly in managerial roles, with accessing and utilizing data from LHIMS, despite its availability. These difficulties stem from a lack of familiarity with basic computer operations, navigation through digital platforms, and understanding the specific functionalities required to manage patient records and health data. This digital literacy gap can also create a sense of frustration or reluctance to engage with the technology, further hindering their ability to adapt to modern healthcare processes that increasingly rely on digital tools.

In Zimbabwe, E-HR was found to significantly enhance decision support for health workers, with features like the e-Partograph automating patient tracking during labor and delivery.

> “*Use of e-Partograph. It has improved so much, the nurse does not have to forget to listen to the foetal heart because it gives alerts and it flicks when the time comes to say now you are supposed to go listen to the foetal heart, to do the vaginal examination, blood pressure*.”[Zim Health Worker]

The integration of clinical practice guidelines within the E-HR system standardized the quality of care and fostered better teamwork through shared patient management histories.

> “*Improved on communication since it allows colleagues to know how I have managed the patient and what further services are needed*” [Zim Health Worker]

However, a notable challenge was the lack of doctor participation in entering patient records, which diminished the overall effectiveness of the system.

> “*I cannot say it has improved. When we capture the observations or vitals when we refer the patient to the doctor, most of the time, they don’t input the data in the system, so there is a missing link.*” [Zim Health Worker]

### Medicines and Supply

Zimbabwe’s E-HR system, through its Pharmacy and Commodity Tracking module, provided real-time tracking of medicines, including stock levels, usage, and expiration dates, and monitored minimum and maximum stock levels.

> “*It shows a breakdown of medicines that are in short supply. Rarely do you miss to order medicines. It tells us that we have, for example we have 7 sachets of paracetamol in stock but only 2 have not expired*” [Zim Nurse Manager]

While this improved inventory management, stock-outs persisted due to broader supply chain challenges that were beyond the scope of the E-HR system.

> “*We order from National Pharmacy (Nat Pharm), but the deliveries come incomplete or with shortages or no deliveries at all*” [Zim Nurse Manager]

Similar issues were noted in Ethiopia, where DHIS2 improved resource allocation but was hampered by infrastructure and technical capacity limitations characterized by high staff turnover, and poor stakeholder coordination.

### Health Care Financing

Findings in the domain health financing and leadership were less explored in all three countries. However, all three countries reported findings on the use of DHIs in resource tracking and allocation, an indirect contribution towards efficient utilisation of resources and hence cost savings. Zimbabwe and Ethiopia noted that despite the potential of DHIs in resource tracking and allocation, limited funding in the health system will continue to result in resource shortages such as continued drug stock outs (Zimbabwe), and high staff turnover (Zimbabwe and Ethiopia).

### Cross-Cutting Issues

All three countries faced significant challenges in DHI implementation, particularly related to infrastructure. In Ethiopia, Ghana and Zimbabwe, frequent power outages, unreliable internet connectivity, and the theft of solar panels (Zimbabwe) and tablets (Zimbabwe) disrupted the functionality of the DHIs.

> “*Here the challenges are no electricity, solar panels stolen, Wi-Fi never worked here because there is no power. E-HR has its own problems which are not common in routine practices*” [Zim Nurse Manager]

The use of personal devices to access LHIMS poses a major security issue in Ghana. Likewise, multiple individuals can have unrestricted access to these personal devices, raising questions about data security and confidentiality.

Key informants highlighted the need for a more comprehensive systems infrastructure to adequately support the implementation of E-HR in Zimbabwe.

> *The existing infrastructure does not support the new system well."*[Zim Key Informant]

Furthermore Ethiopia, noted the infrastructure in many rural and remote areas remains inadequate, with unreliable internet connections and frequent power outages disrupting the functionality of the system. These challenges hinder the full utilization of DHIS2, preventing healthcare providers from accessing or updating information in real-time. The effectiveness of the system was thus compromised, as healthcare workers were often unable to enter data or generate reports promptly, leading to delays in decision-making.

In Ghana, that older healthcare professionals, often in managerial positions, struggled with retrieving information from LHIMS, despite it being readily available for decision-making. The ease of accessing information was identified as a contributing factor to delays in data utilization by PHCs in Ghana. For Zimbabwe integration of E-HR in clinical workflows particularly within high volume sites was a challenge. Ethiopia noted DHIS 2 challenges related to data quality and interoperability. In some cases, the data entered into DHIS2 may be incomplete, inaccurate, or inconsistent, which undermines its reliability for decision-making. There are also concerns about how well DHIS2 interacts with other health information systems, both within Ethiopia and at the regional level. Interoperability problems make it difficult to consolidate data from multiple sources, hindering the integration of health information across various platforms and preventing a comprehensive view of the healthcare system. For Ghana, the dual use of DHIMS and LHIMS without interoperability of these two systems has resulted in duplication and data inaccuracies. In Zimbabwe, although E-HR is interoperable with DHIS2, the reliance on paper record systems as a backup during E-HR downtime has led to inconsistent and inaccurate data. This is primarily due to the high human resource costs associated with updating historical data into the electronic system once it is operational again The sustainability of DHIS2 was a significant concern in Ethiopia. The system’s long-term success depended on adequate financial resources, continued government commitment, and ongoing support from development partners.

## Discussion

In a scoping review of Digital Health Interventions and Health Systems Strengthening from sub-saharan Africa(4), the overwhelming representation of health services (82%; n=603 out of the 738 DHIs included) was mentioned whereas leadership and governance aspect of health systems was least represented in the DHI literature (0.4%;n = 3). Thirty-four percent (34%, n = 252) of the digital interventions strengthened more than one of the six HSS building blocks. The largest proportion (84%) were focused on mining data, as opposed to improving provision of services. Our paper explored how established DHIs in the three countries contributed towards more than one of the health systems building blocks. Our study investigated health information management system DHIS2 for health data reporting and patient management information systems (LHMIS in Ghana and Impilo E-HR in Zimbabwe) for their contribution towards improving services across the 6 health systems building blocks. Both systems for health data reporting and patient management DHIs are widely implemented and established in the three countries studied. All countries use DHIS2 for health data reporting, with implementation spanning from primary care facilities to the national level, providing performance indicator data at each stage of care. According to the WHO Digital Health Atlas (DHA), DHIS2 is the most widely adopted DHI in Africa, with 56% of all DHIS2 implementations in Africa (9). Of the three countries, only in Zimbabwe is DHIS2 integrated with the electronic health record system, enhancing efficiencies in health data management and enabling real-time reporting. There are notable contributions of these DHIs to health systems strengthening. These DHIs are now established and their contributions to health systems strengthening acknowledged across all service points in the primary health care system.

### Service Delivery

DHIS 2 implementation in Ethiopia was noted to improve service delivery by increasing accurate information availability for improved service delivery. In Zimbabwe, E-HR streamlined clinical workflows, and increased efficiency of service delivery. This is facilitated by increasing information availability, sharing at point of decision making (4). E-HR users also noted the reduced fragmentation of information across different caregivers and different service points, consolidating the information available for a holistic response at the point of care, a finding that has been noted by (10). The integrated decision support tools, also strengthened service delivery, enhancing the quality of care at primary care. The use of tools such as the incorporation of clinical practice guidelines and services like ePartograph have standardised the care offered by caregivers. However, the challenges of managing high patient volumes negatively affect service delivery, with slow response times at the point of care discouraging the use of electronic health records (E-HR) during patient interactions. To address this, E-HR solution developers should focus on implementing decentralized solutions where data is locally available and synchronized during off-peak hours. Some strategies to consider include:

1. **Local Data Storage and Processing**: Deploying localized servers at healthcare facilities to securely store and process patient data, particularly for frequently accessed cases. This approach ensures faster data retrieval and minimizes dependency on centralized systems, thereby enhancing system responsiveness and reliability during peak usage periods.
2. **Offline Functionality with Scheduled Synchronization**: Developing E-HR systems that allow healthcare providers to access and update patient information offline. The data can then be automatically synchronized with central servers during off-peak hours, minimizing network load during high-traffic periods.

### Health Information

All three country studies reported improvements in health information systems within the health systems building blocks. Primary and secondary healthcare facilities have widely adopted digital health interventions (DHIs) across the continuum of care. Key improvements have been noted in data quality—specifically in terms of completeness, timeliness, and accuracy—as well as in data availability, accessibility, and accountability. At the point of care, electronic health record (E-HR) systems have enhanced the quality of patient records by incorporating audit trails to track changes and implementing integrity rules to ensure data accuracy. The integration of E-HR with DHIS 2 has significantly reduced the administrative burden of updating paper records and increased the availability of real-time data. In Ghana, both the District Health Information Management System (DHIMS) and Local Health Information Management System (LHIMS) have played a crucial role in enabling data-driven decision-making for primary healthcare (PHC) managers, particularly in forecasting, monitoring, and evaluating performance. These DHI capabilities serve as catalysts for advancing Learning Health Management Information Systems, which evolve through the progressive maturation of data use at individual, team, and organizational level ultimately fostering adaptive and responsive health systems (11).

The lack of integration between these systems has led to data duplication and inaccuracies. Therefore, systems integration is a critical component to strengthen between reporting and patient care information systems.

### Health Workforce

Health workers have widely embraced DHIs at the point of care and for reporting. In Ethiopia, policy makers have adopted the use of DHIS 2 data for real time decision making. In Ghana, 88% of PHC managers rely on LHIMS data for administrative and managerial functions. In Zimbabwe, all nurse managers and nurse caregivers use E-HR in their daily practices. However, more efforts are needed to ensure inclusivity, particularly among older healthcare professionals who face challenges in adopting these tools. Some of these strategies can include:

### Targeted Training and Mentorship

To improve adoption among older healthcare professionals and policy makers, focused training, mentorship, and ongoing refresher courses should be provided. These programs can be designed to address their specific concerns and learning needs, ensuring they feel confident in using digital tools in their workflow.

### Integration of DHIs in Health Training Curriculum

Incorporate E-HR training into medical school curricula to ensure that future doctors are equipped with the necessary skills to use these systems from the start of their careers.

### Integrate DHIs into Standard Operating Procedures

Review program implementation and patient care to incorporate DHIs as a routine part of care. Clearly define the roles of various healthcare cadres and identify where each DHI fits within the continuum of care hierarchy. This integration should ensure that DHIs become a standard element of health service delivery.

### Medicines and Supply

DHIs have been recognized for their positive impact on resource tracking. In Ethiopia and Ghana, policymakers highlighted the utility of DHIS 2 in tracking commodity resources at the policy level. In Zimbabwe, electronic health records (E-HRs) have strengthened commodity tracking at the facility level, enhancing accountability for medicines and promoting more efficient use of drugs. This has resulted in a reduction in commodity losses due to expiration and pilferage.

However, despite these improvements, DHIs alone are insufficient. For their full potential to be realized, they must be supported by broader efforts to strengthen other health system building blocks. The persistence of stockouts, driven by the underfunding of essential commodities, underscores the need for better resource allocation and overall system strengthening.

### Health Financing

This was not explored extensively in the studies. However, the improvements in resource utilization have indirectly benefited healthcare financing by enhancing efficiency and reducing costs associated with resource wastage and system leakages. Therefore, further research is needed to better understand how DHIs contribute to the health financing building block and their overall impact on cost-effectiveness in healthcare systems.

### Cross-cutting Issues

The implementation of DHIs is accompanied by several challenges that are common across the three countries. The existing infrastructure is insufficient to fully support DHIs, with power and internet connectivity at facility and district levels being critical weaknesses. Additionally, high staff attrition in the health sector negatively impacts the effective use of these systems, as trained personnel frequently leave, creating gaps in skilled users. Furthermore, the lack of interoperability between different systems introduces challenges such as redundancy, data duplication, and compromised data quality.

## Conclusion

These studies underscore the potential of DHIs in strengthening health systems in resource-limited settings. The cases of Ethiopia, Ghana, and Zimbabwe demonstrate that DHIs, such as DHIS2, LHIMS, and the *Impilo* EHR system, significantly enhance data management, accountability, and decision-making. These systems contribute to improved service delivery, resource allocation, and healthcare equity. However, critical challenges, including infrastructure deficiencies, technical capacity gaps, high staff turnover, and interoperability limitations, continue to hinder the full realization of their benefits.

## Data Availability

The data is largely qualitative interviews in Excel Sheets. These can be made available online as requested

## Acknowledgements

This work received financial support from the WHO Alliance for Health Policy and Systems Research (Alliance). The Alliance is able to conduct its work thanks to the commitment and support from a variety of funders. These include our long-term support from the Swedish International Development Cooperation Agency (SIDA) and the Norwegian Agency for Development Cooperation (NORAD), as well as designated funding for specific projects within our current priorities. For the full list of Alliance donors, please visit https://ahpsr.who.int/about-us/funders.

